# Analysis of R-loop forming regions identifies *RNU2-2P* and *RNU5B-1* as neurodevelopmental disorder genes

**DOI:** 10.1101/2024.10.04.24314692

**Authors:** Adam Jackson, Nishi Thaker, Alexander Blakes, Siddharth Banka

## Abstract

R-loops are critical for gene regulation but their contribution to human Mendelian disorders has not been explored. We show that in rare disease cohorts *de novo* variants are highly prevalent in genomic regions which form R-loops. Using this insight, we demonstrate that variants in two novel disorder genes, *RNU2-2P* and *RNU5B-1*, together with the recently described *RNU4-2* provide a genetic explanation for an exceptionally large proportion of individuals with suspected rare Mendelian disorders.

## Main

R-loops are DNA-RNA hybrid structures, which form at sites of active transcription.^1^ These structures mark transcribed regions and also play important roles in promoter DNA hypomethylation,^2^ histone modification^3^ and DNA replication.^4^ Even though genomic regions that form R-loops (henceforth, R-loop regions) are functionally important, germline variants in them have not been systematically explored in the context of human diseases.

In 13,949 trios in the 100,000 Genomes Project (100KGP, Genomics England, The National Genomics Research Library v5.1, Genomics England. doi:10.6084/m9.figshare.4530893/7. 2020) database,^5^ we identified 53,342 (median 4 per trio, range 1 - 46) de novo (DN) variants occurring within experimentally-determined R-loop regions.^6^ We found comparable frequency of DN R-loop region variants (RRVs) in a whole genome sequencing cohort of 1,548 healthy trios,^7^ but DN RRVs in snRNA genes were relatively enriched the disease cohort (**Figure 1A**) (**Supp Table S1**).

**Figure 1.**
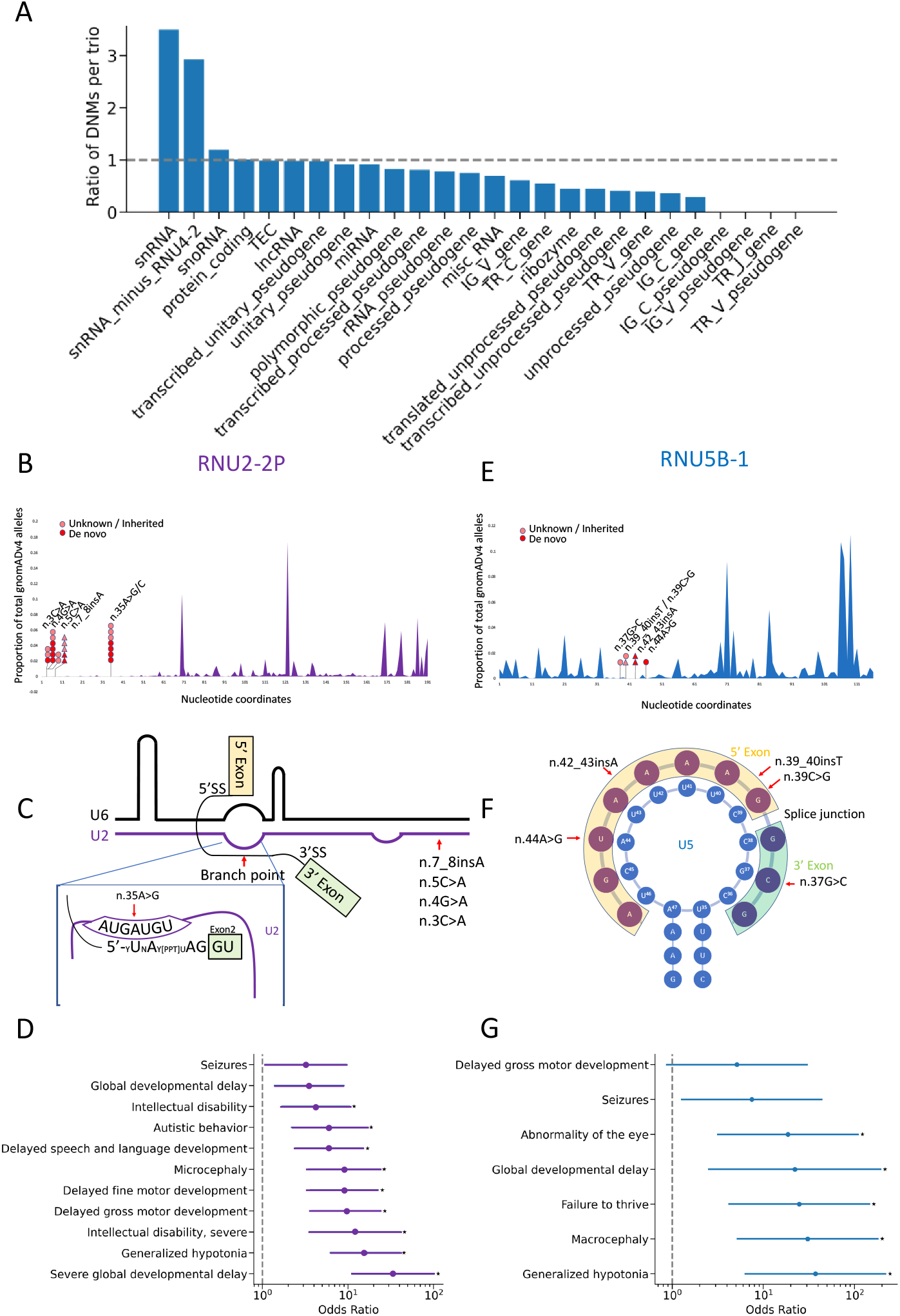
(A) Bar plot of the ratio of DN mutations in 100KGP versus the Icelandic cohort in different GENCODE gene types (B) Allele frequences of *RNU2-2P* variants in gnomAD v4.1. SNVs (filled circles) and indels (filled triangles) in affected patients are shown. (C) Schematic representation of RNU2-2P variants mapped to the U2-U6 structure in complex with the mRNA branch point. (D) Odds ratios for Human Phenotype Ontology terms in RNU2-2P cases compared to all probands in the rare disease arm of 100KGP. Only HPO terms observed in at least three *RNU2-2P* cases are shown. Asterisks indicate phenotypes which are significantly over-represented (two-sided Z test of the log odds ratio, Bonferroni p<0.0045) (E) Allele frequencies of *RNU5B-1* variants gnomAD v4.1. SNVs (filled circles) and indels (filled triangles) in affected patients are shown. (F) Schematic representation of *RNU5B-1* variants mapped to the U5 structure in complex with the acceptor and donor sites of adjacent exons, amended from Artemyeva-Isman et al.^18^ (G) Enrichment analysis of Human Phenotype Ontology terms in *RNU5B-1* cases compared to all probands in the rare disease arm of 100KGP. Only HPO terms observed in at least two *RNU5B-1* cases are shown. Asterisks indicate phenotypes which are significantly over-represented (two-sided Z test of the log odds ratio, Bonferroni p<0.0071)

To identify potentially disease causing DN RRVs, we prioritised variants present in three or more probands but absent from controls in the 100KGP dataset. This revealed eight candidate RRVs, which included the recently described highly recurrent pathogenic indel (ENST00000365668.1:n.64_65insT) in *RNU4-2*^8,9^ in 31 individuals, four other known pathogenic variants in 13 participants and one variant of uncertain significance **(Supp Table S2)**. Excluding *RNU4-2* from the analysis revealed that DN RRVs in snRNA genes were still relatively enriched the disease cohort suggesting additional disease related variants (**Figure 1A**). We identified six individuals with two recurrent DN RRVs in another gene, *RNU2-2P* (ENST00000410396.1:n.4G>A and n.35A>G). *RNU2-2P* is annotated as a pseudogene but encodes a U2 small nuclear RNA that is incorporated into the spliceosome^10^ and is expressed in the cerebral hemispheres (**Supp Figure 1**). Somatic *RNU2-2P* variants are found in multiple cancers.^11^ Both variants are present in a highly constrained 5’ region of *RNU2-2P* (from 1 to 60bp) (**Figure 1B**). Within this constrained region, we detected four more individuals with additional DN variants (n.3C>A, n.35A>C and n.7_8insA) in the 100KGP dataset. Six other individuals were also found to carry one of these five variants in heterozygous state (parental samples not available in four cases, one instance each of maternal and paternal transmission) in the 100KGP database. We acknowledge similar results in an independent study^12^. Analysis of the Genomics England National Genomics Research Library (NGRL) and SolveRD^8^ (predominantly exome sequencing) databases revealed five more individuals with heterozygous *RNU2-2P* variants affecting the constrained region. Variant positions n.3, n.4, n.5 and n.7 are located within the 5’ end of U2, which directly interacts with the 3’ end of U6 in the major spliceosome (**Figure 1C**).^15^ The n.35A nucleotide is in the branch site recognition sequence (GU<u>A</u>GUA) that base-pairs with the conserved U of the branch point (YNY<u>U</u>RAY, where Y, pyrimidine; N, any nucleotide; and R, purine).^16^ The pathogenic n.35A>G variant we report is predicted to replace A_3_-U_4_ with G_3_-U_4_ wobble base-pairing. Interestingly, in ∼2% of branch points U_4_ is replaced by C_4;_^16^ n.35A>G could lead to stronger G^3^-C^4^ pairing at these loci (**Supp Figure 3**). Interrogation of the phenotype of the 18 individuals with *RNU2-2P* variants in 100KGP showed that all were genetically unsolved and their significantly overrepresented phenotypes included severe global developmental delay (OR 33.6, p<10^−8^), generalised hypotonia (OR 15.8, p=1.2×10^−*8*^) and microcephaly (OR 9.0, p=1.1×10^−5^) (**Figure 1D**). Variant information for all 23 individuals with *RNU2-2P* variants is provided in **Supp Table S3**, whilst detailed clinical information on two individuals from our centre are provided as Supplementary Case Reports (**Supp Figure 4**).

Based on the high frequencies of *RNU4-2* and *RNU2-2P* and the relative enrichment of DN RRVs in snRNAs in the disease cohort (**Figure 1A**), we relaxed our filtering strategy for DN RRVs overlapping with snRNAs to include recurrent variants present in only two probands and removed the high stringency criteria for DN variant detection. This led to the identification of a recurrent DN variant in *RNU5B-1* (ENSG00000200156.1:n.42_43insA) in two individuals. Manual inspection of read alignments confirmed the DN origin of this variant in both cases. This variant occurs within a 20bp region deplete of variation in gnomADv4 (**Figure 1E**), which corresponds to the loop I structure of U5 essential for recognising both acceptor and donor splice sites in apposition (**Figure 1F**).^17,18^ Expansion of the search for other variants within this critical region of *RNU5B-1* revealed three other individuals in the 100KGP with three different variants (n.37G>C and n.39_40insT of unknown inheritance, and a DN n.44A>G change). All five individuals were genetically unsolved, and had consistent phenotypes, among which generalised hypotonia (OR 37.3 p=7.4×10^−5^), macrocephaly (OR 30.6, p=1.8×10^−4^), failure to thrive (OR 24.7 p=4.5×10^−4^), global developmental delay (OR 22.1 p=5.6×10^−3^) and abnormality of the eye (OR 18.7 p=1.4*10^−3^) were significantly over-represented versus other rare disease probands in the cohort (**Figure 1G**). In the NGRL cohort, we identified one more unsolved individual with NDD and macrocephaly and n.39C>G (variant not inherited maternally). Notably, n.39C>T is observed in 3 individuals in gnomADv4 and is likely tolerated because U can wobble base-pair with the final G of the 5’ exon,^19^ whereas other substitutions are predicted to abolish binding. Variant information for all six individuals with *RNU5B-1* variants is provided in **Supp Table S4**.

In summary, we show that DN variants are highly prevalent in genomic regions which form R-loops and variants in snRNA components of the major spliceosome show particular enrichment in a disease cohort in comparison with a control cohort. We implicate rare variants in *RNU2-2P* and *RNU5B-1* as causes for highly frequent novel NDDs. *RNU2-2P*-related disorder is characterised by global developmental delay, intellectual disability, microcephaly microcephaly, autistic behaviour and tendency for seizures. *RNU5B-1*-related disorder is characterised by global developmental delay, hypotonia, macrocephaly and failure to thrive (**Supp Figure 5**). Collectively, they provide a genetic explanation for exceptionally large proportion of individuals with suspected rare Mendelian disorders, accounting for the diagnoses of at least 0.54% of individuals with NDDs in the 100KGP project.

## Data Availability

All data used in this study was accessed through the Genomics England Research Environment or Solve-RD RD-CONNECT platform. Access to these datasets can be granted to approved researchers pending review by the appropriate board.

## Online Methods

### Project Registration and Data Access

The project was registered with the Genomics England research registry (RR1147) and received approval to access data from 100KGP ((Genomics England, The National Genomics Research Library v5.1, Genomics England. doi:10.6084/m9.figshare.4530893/7. 2020.). The consensus RL region’s BED file was imported to the Genomics England Research Environment (GERE) using the Airlock and all patient data was exported through the Airlock for preparation of this manuscript.

### Identification and Characterization of De Novo Variants within R-loop regions using 100,000 Genomes Project

We used the consensus R-loop regions (RL regions) provided by Miller et al 2022 derived from 810 R-loop mapping datasets in humans were used. Chromosomal coordinates, aligned to GRCh38, were downloaded from the supplementary data and converted to BED format. We then intersected this BED file with a BED file generated from the DN variant dataset in GERE (main_programme_v18_2023-12-21). Intersection was performed using the bedtools intersect function, with default parameters. GRCh37 coordinates in the GERE were lifted over to GRCh38 using UCSC-liftOver. We included high confidence DN variants in GERE flagged by a stringency criteria of ‘1.’ For the relaxed analysis of DN RRVs in snRNAs, we included DN variants which failed the stringency filter but were validated by manual inspection of sequencing reads in IGV

### Analysis of DNM occurring in Icelandic control dataset versus 100KGP

We downloaded the DNM file for the Icelandic study and generated a BED file of coordinates. We then intersected these with the GENCODE v32 GTF file and extracted overlaps with genes and then counted the number of overlaps for each gene type in GENCODE. We repeated this analysis for DNM in 100KGP. We used the genomic footprint of each gene type to calculate the number of DNM per bp per trio for each group. We then calculated the ratio of DNMs comparing 100KGP to Icelandic controls.

### Filtering and Prioritization of Variants

DNMs overlapping RL regions were then subject to filtering based on their allele frequency in gnomADv3 and also their allele frequency within the 100KGP aggregated dataset (aggV2), encompassing data from 78,195 participants. Using aggV2, we filtered for variants that were absent from gnomADv3, occurred exclusively in probands in the 100KGP and affected over three participants with at least three DN occurrences. The number of DN occurrences was calculated using the DNM dataset using stringency ‘1’ variants only.

### Phenotype Matching and Analysis

Phenotypic information for individuals carrying candidate variants was extracted from the LabKey rare disease phenotype database (main_programme_v18_2023-12-21) using HPO terms. The analysis focused on identifying common phenotypes across individuals sharing these variants. For individuals with *RNU2-2P* and *RNU5B-1* variants. we counted the occurrence of each HPO term and calculated the odds ratios for these terms against all other individuals in the cohort. Significance thresholds were adjusted by Bonferroni correction.

HPO terms for individuals with variants in *RNU4-2* were taken from the supplementary information of the recent publication describing the disorder.^8^

### Analysis of constraint in gnomADv4

Variants within the *RNU2-2P* transcribed region were downloaded from the gnomADv4 browser. Allele counts were combined for multiallelic variants. The fraction of all *RNU2-2P* variants in gnomADv4 was then calculated by dividing the nucleotide pooled variant alleles by the total number of variant alleles.

### Structural analysis of RNU2

Candidate variants in *RNU2-2P* were annotated onto the crystal structure of U2 in complex with MINX mRNA branch point. *RNU2-1* and *RNU2-2P* have 95.8% identify and a completely conserved GUAGUA branch point binding motif. The Protein Data Bank Mol* (WebGL) server was used to navigate the structure and images were downloaded and annotated in PowerPoint.

Models of wild-type and mutant *RNU2-2P* were generated using the AlphaFold3 web server. As input, we used GUAGUA as the wild-type fasta sequence for U2 and UACUAAC as the canonical branch point. The first model generated (_0) in AlphaFold3 was then used for each prediction and subsequent analysis in PyMol.

### Calculation of the proportion of unexplained NDD caused by RRVs

We used the calculations by Chen et al^8^ and added *RNU2-2P* and *RNU5B-1* cases. This meant a calculation of 80 individuals affected by variants in *RNU4-2, RNU2-2P* or *RNU5B-1* divided by 14735 (the calculated number of unsolved cases in 100KGP using 40% as the diagnostic yield).

## Supplementary Material

### Supplementary Case Reports

**Supplementary Table S1:** Table showing the number of trios examined and the number of DN variants per trio in both control and disease cohorts as well as the overlaps with R-loop regions

**Supplementary Table S2:** All candidate DN variants occurring within R-Loop regions in the 100KGP passing stringent filters for gnomADv3 allele frequency and present in at least three probands with de novo origin. Multi-allelic candidate variants were removed if at least one allele at the position was present in gnomADv4.

**Supplementary Table S3:** Variant nomenclature for all individuals with *RNU2-2P* variants in this study.

**Supplementary Table S4:** Variant nomenclature for all individuals with *RNU5B-1* variants in this study.

**Supplementary Figure 1:**
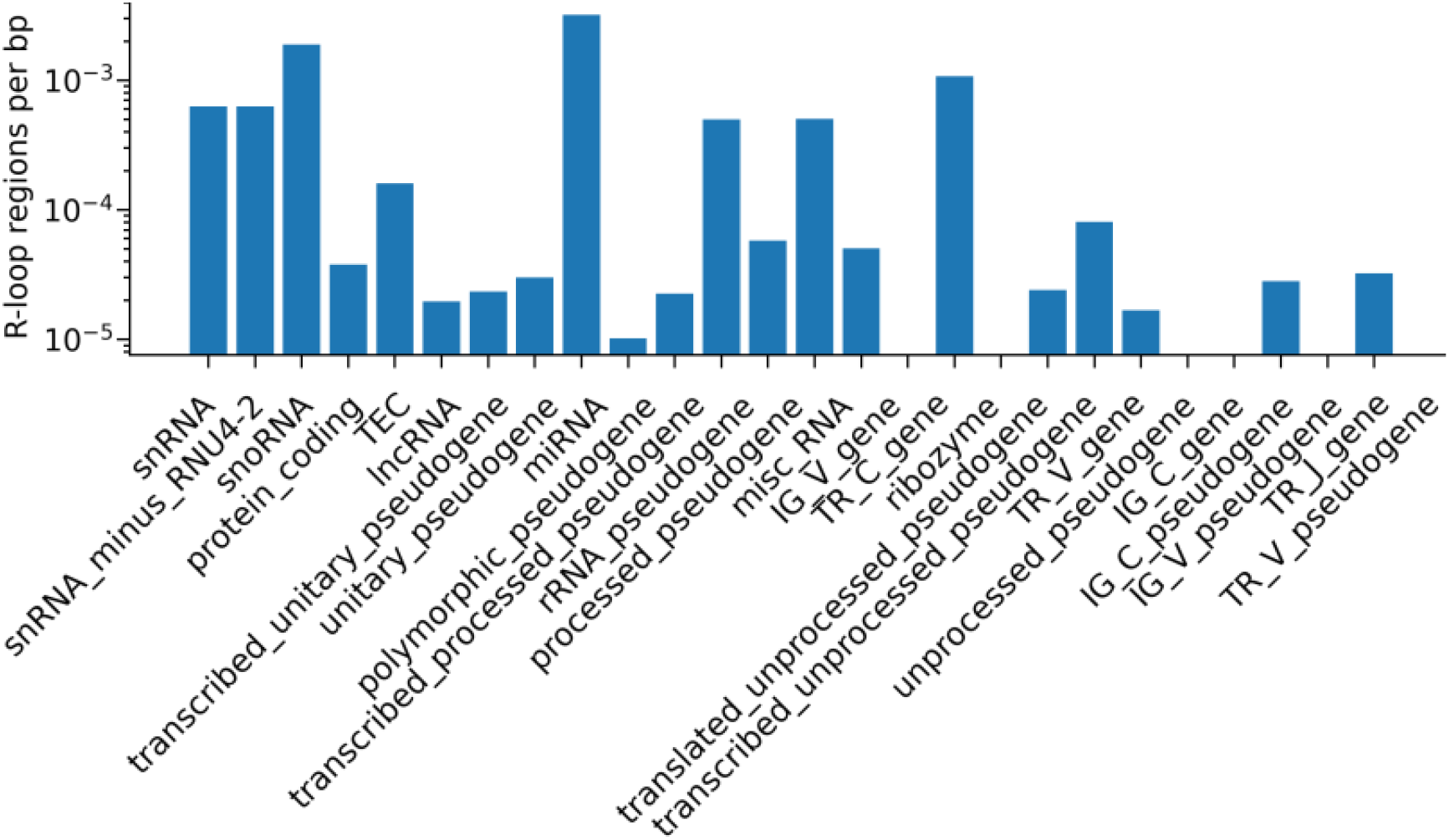
Bar plot of R-loop density in different GENCODE gene types.

**Supplementary Figure 2:**
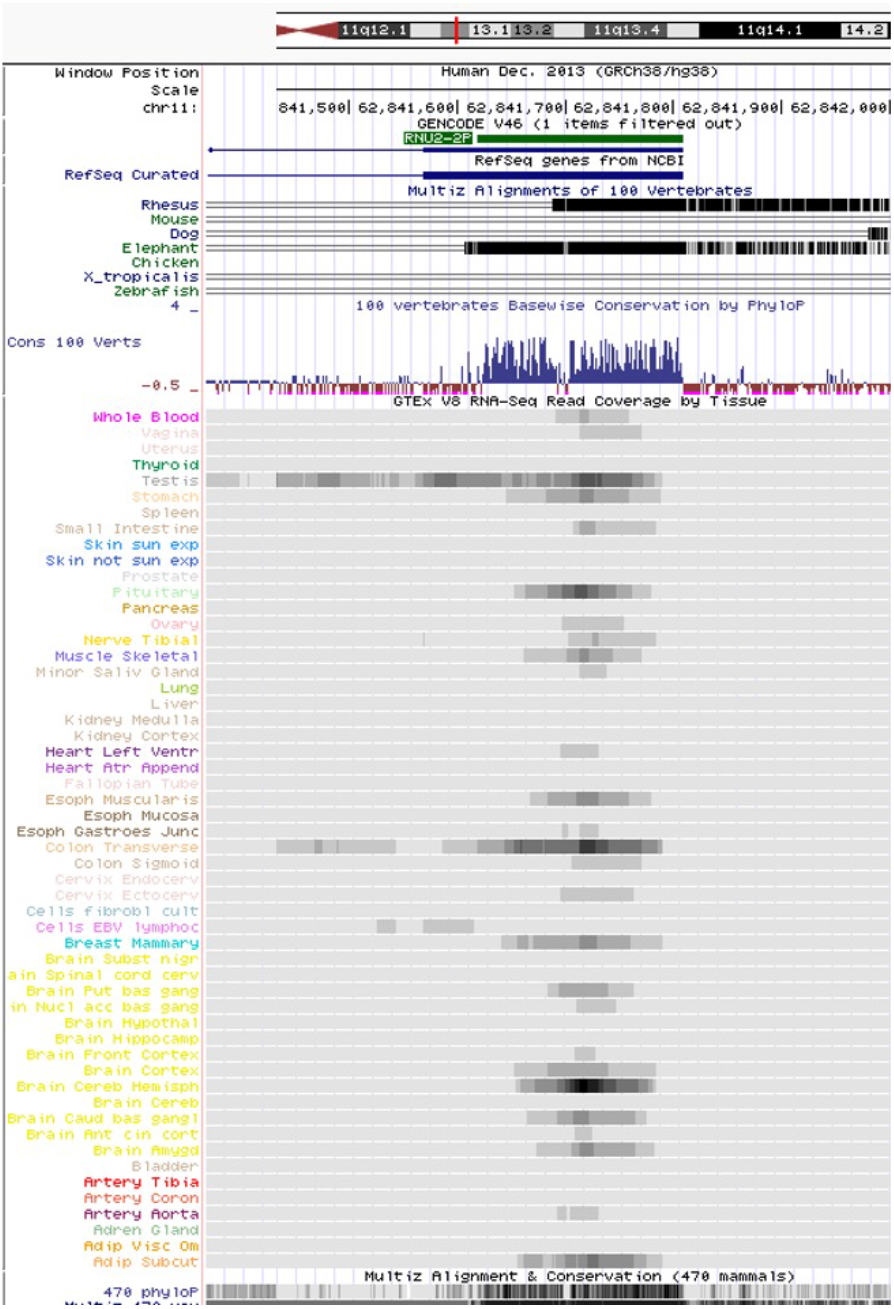
UCSC screenshot of GTEx coverage for RNU2-2P as well as evolutionary conservation.

**Supplementary Figure 3:**
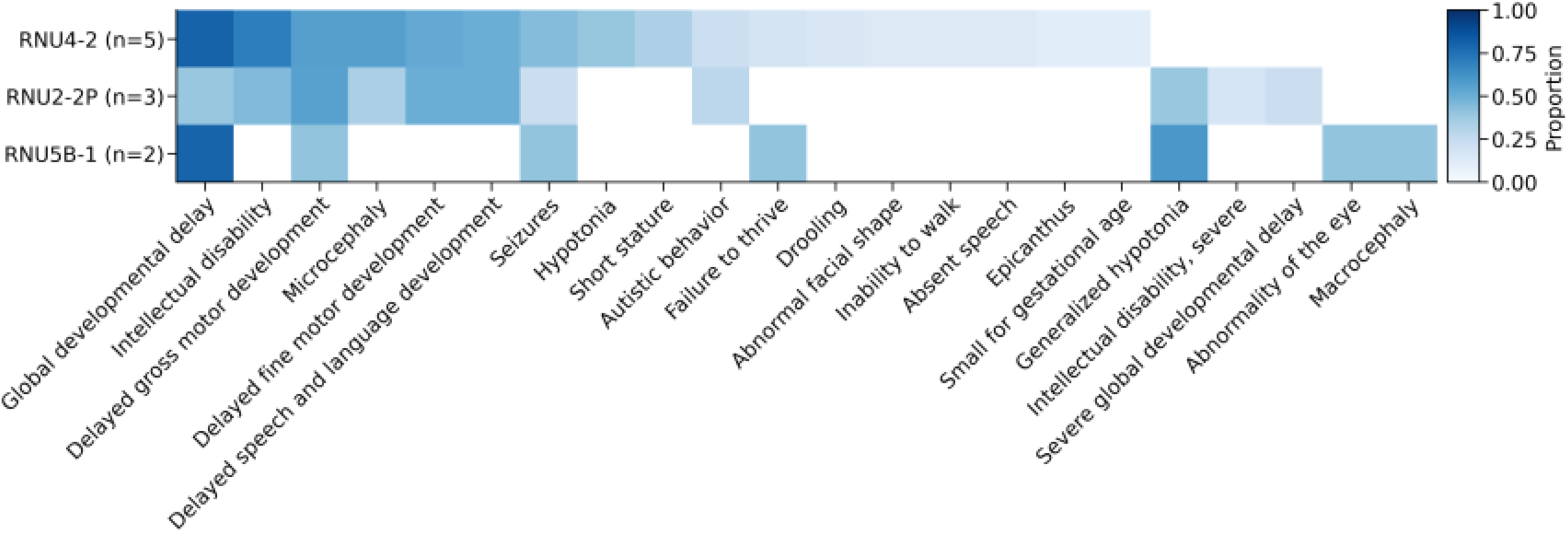
AlphaFold predictions of interactions between U2 n.35 in both wild-type (A, top) and mutant (G, bottom) constitutions with consensus branch points with U^4^ (left) and alternative branch points with C^4^ (right)

**Supplementary Figure 4:**
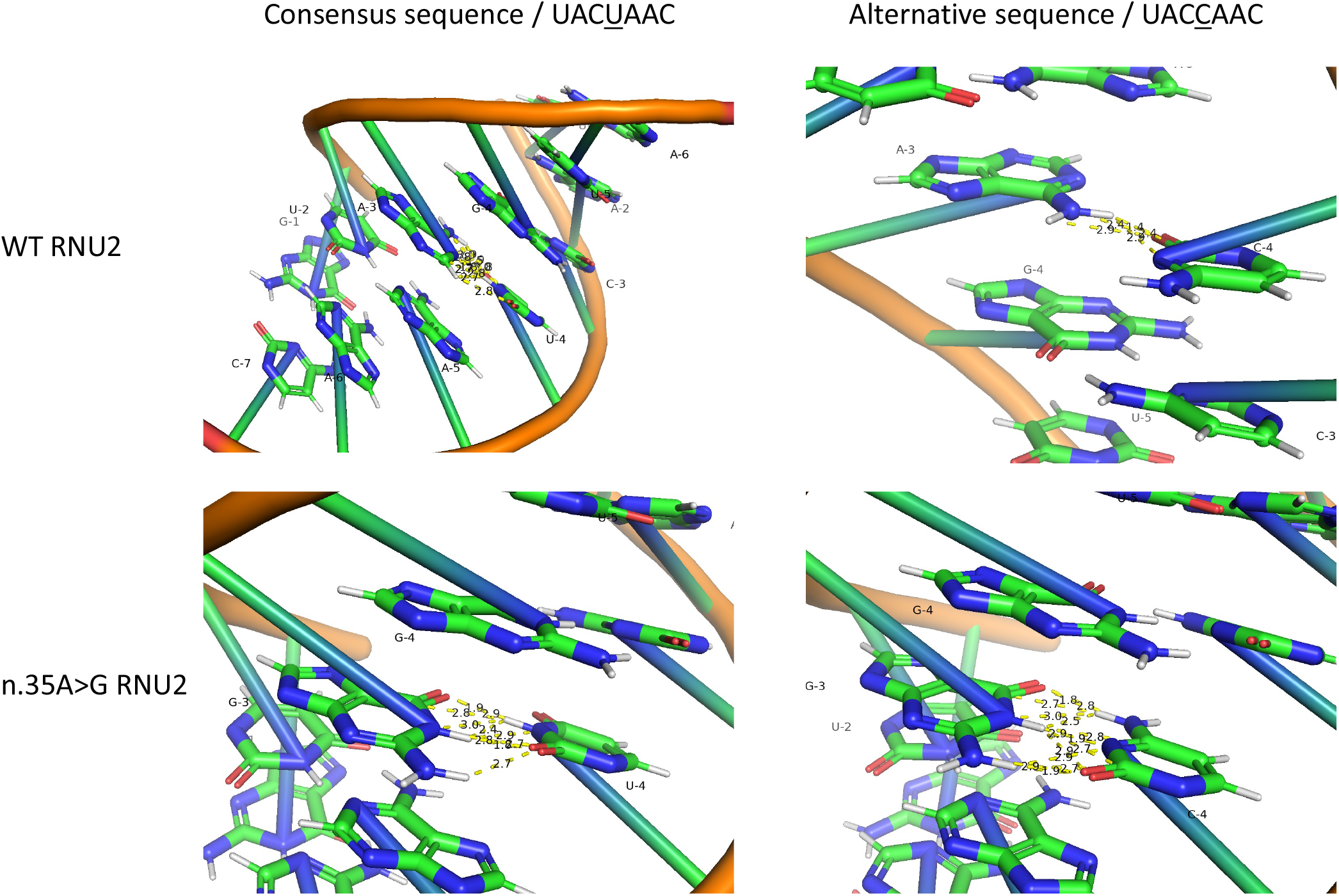
(A) Facial photographs of Individuals I (*RNU2-2P*: DN n.3C>A) and Individual II (*RNU2-2P*: DN n.4G>A). (B) T2-weighted MRI brain images from 4 and 10 years of Individual II.

**Supplementary Figure 5:** Heatmap demonstrating overlapping and distinct HPO terms in *RNU4-2, RNU2-2P* and *RNU5B-1* disorders. For each disorder, only HPO terms present in at least ‘n’ individuals are represented due to the different sample sizes.

## Acknowledgments

We thank the participants of the 100,000 Genomes Project. We thank Genomics England for generating the data and provided the GERE platform for analysis. Thanks to Peter O’Donovan, Zainab Mustafa, Ana Lisa Taylor and Joanne Yang for airlock requests. A.J. and S.B. acknowledge the support of Solve-RD. The Solve-RD project has received funding from the European Union’s Horizon 2020 research and innovation program under grant agreement number 779257. This study has been delivered through the National Institute for Health and Care Research (NIHR) Manchester Biomedical Research Centre (BRC) (NIHR203308). The views expressed are those of the author(s) and not necessarily those of the Solve-RD, the NIHR or the Department of Health and Social Care. AB is supported by a Wellcome PhD Training Fellowship for Clinicians and the 4Ward North PhD Programme for Health Professionals (223521/Z/21/Z). This research was made possible through access to data in the National Genomic Research Library, which is managed by Genomics England Limited (a wholly owned company of the Department of Health and Social Care). The National Genomic Research Library holds data provided by patients and collected by the NHS as part of their care and data collected as part of their participation in research. The National Genomic Research Library is funded by the National Institute for Health Research and NHS England. The Wellcome Trust, Cancer Research UK and the Medical Research Council have also funded research infrastructure.

## Author contributions

NT, AJ and AB performed analyses. AJ and SB wrote the manuscript and supervised the project. All authors have read and approved the submitted manuscript.

## References

1. Ariel, F. et al. R-Loop Mediated trans Action of the APOLO Long Noncoding RNA. Molecular Cell 77, 1055-1065.e4 (2020).

2. Ginno, P. A., Lott, P. L., Christensen, H. C., Korf, I. & Chédin, F. R-Loop Formation Is a Distinctive Characteristic of Unmethylated Human CpG Island Promoters. Molecular Cell 45, 814–825 (2012).

3. Li, H. et al. The Cumulative Formation of R-loop Interacts with Histone Modifications to Shape Cell Reprogramming. Int J Mol Sci 23, 1567 (2022).

4. Lombraña, R., Almeida, R., Álvarez, A. & Gómez, M. R-loops and initiation of DNA replication in human cells: a missing link? Front. Genet. 6, (2015).

5. The 100,000 Genomes Project Pilot Investigators. 100,000 Genomes Pilot on Rare-Disease Diagnosis in Health Care — Preliminary Report. New England Journal of Medicine 385, 1868–1880 (2021).

6. Miller, H. E. et al. Quality-controlled R-loop meta-analysis reveals the characteristics of R-loop consensus regions. Nucleic Acids Res 50, 7260–7286 (2022).

7. Jónsson, H. et al. Parental influence on human germline de novo mutations in 1,548 trios from Iceland. Nature 549, 519–522 (2017).

8. Chen, Y. et al. De novo variants in the RNU4-2 snRNA cause a frequent neurodevelopmental syndrome. Nature 632, 832–840 (2024).

9. Greene, D. et al. Mutations in the U4 snRNA gene RNU4-2 cause one of the most prevalent monogenic neurodevelopmental disorders. Nat Med 30, 2165–2169 (2024).

10. Mabin, J. W., Lewis, P. W., Brow, D. A. & Dvinge, H. Human spliceosomal snRNA sequence variants generate variant spliceosomes. RNA 27, 1186–1203 (2021).

11. PanCancer analysis of somatic mutations in repetitive regions reveals recurrent mutations in snRNA U2 - PMC. https://www.ncbi.nlm.nih.gov/pmc/articles/PMC8921233/.

12. Greene, D. et al. Mutations in the U2 snRNA gene RNU2-2P cause a severe neurodevelopmental disorder with prominent epilepsy. 2024.09.03.24312863 Preprint at 10.1101/2024.09.03.24312863 (2024).

13. Matalonga, L. et al. Solving patients with rare diseases through programmatic reanalysis of genome-phenome data. Eur J Hum Genet 29, 1337–1347 (2021).

14. Zurek, B. et al. Solve-RD: systematic pan-European data sharing and collaborative analysis to solve rare diseases. Eur J Hum Genet 29, 1325–1331 (2021).

15. Rhode, B. M., Hartmuth, K., Westhof, E. & Lührmann, R. Proximity of conserved U6 and U2 snRNA elements to the 5′ splice site region in activated spliceosomes. The EMBO Journal 25, 2475–2486 (2006).

16. Kadri, N. K., Mapel, X. M. & Pausch, H. The intronic branch point sequence is under strong evolutionary constraint in the bovine and human genome. Commun Biol 4, 1–13 (2021).

17. McGrail, J. C., Tatum, E. M. & O’Keefe, R. T. Mutation in the U2 snRNA influences exon interactions of U5 snRNA loop 1 during pre-mRNA splicing. The EMBO Journal 25, 3813–3822 (2006).

18. Artemyeva-Isman, O. V. & Porter, A. C. G. U5 snRNA Interactions With Exons Ensure Splicing Precision. Front. Genet. 12, (2021).

19. Varani, G. & McClain, W. H. The G·U wobble base pair. EMBO reports 1, 18–23 (2000).

